# Altered amyloid plasma profile in patients with disabling headaches after SARS-CoV-2 infection and vaccination

**DOI:** 10.1101/2024.10.19.24315794

**Authors:** Anne Hege Aamodt, Thor Ueland, Marion Boldingh, Burcu Ella Bezgal, Maria Bengtson Argren, Cecilia Adele Dunne, Kari Otterdal, Ida Gregersen, Vigdis Bjerkeli, Annika Elisabet Michelsen, Andreas Husøy, Åse Hagen Morsund, Kristina Devik, Anne Christine Poole, Kristine Bodding Gjendemsjø, Katrin Schlüter, Sara Maria Mathisen, Mari Aalstad-Johansen, Thor Håkon Skattør, Julie Sønnervik, Turid Birgitte Boye, Trine Haug Popperud, Einar August Høgestøl, Hanne Flinstad Harbo, Fridtjof Lund-Johansen, Pål Aukrust, Erling Tronvik, Tuva Børresdatter Dahl, Bente Evy Halvorsen

## Abstract

**Background and objectives:** New onset persistent headache has been reported following acute COVID-19 and to some degree also after SARS-CoV-2 vaccination. The mechanisms for these headache types are unclear. The purpose of this study was to assess levels of amyloid related biomarkers in patients with persistent headache after COVID-19 and SARS-CoV-2 vaccine.

**Methods:** In this prospective observational cohort, patients with severe headache as the dominating symptom after COVID-19 (n=29) and SARS-CoV-2 vaccination (n=31), had neurological assessments with reassessments after 6 months. Plasma levels of amyloid precursor protein (APP), pregnancy zone protein (PZP), cathepsin L1 (CTSL) and serum Amyloid A (SAA1) were measured by ELISA in relation to levels in healthy controls (n=16).

**Results:** We found a strong and persistent upregulation of APP in patients with headache after COVID-19 as compared to the two other groups. At both inclusion and after 6 months APP levels were also increased in those with accompanying cognitive symptoms. In contrast, plasma levels of PZP were elevated in patients with headache after SARS-CoV-2 vaccination at both inclusion and after 6 months as compared to healthy controls. CTSL was only elevated in those with COVID-19 associated as compared with those with vaccine associated headache at baseline, whereas SAA1 showed levels comparable in all groups.

**Conclusion:** Altered plasma levels of soluble markers potentially reflecting changes in amyloid processing was found in patients with persistent headache after SARS-CoV-2 vaccine and particular in those with persistent headache after COVID-19 where we also found some association with cognitive symptoms.

NCT04576351

NCT05235776

**What is already known on this topic:** New onset persistent headache occurs in a subset of individuals after COVID-19 and to some extent after SARS-CoV-2 vaccine. However, the pathophysiological mechanisms are unknown.

**What this study adds:** Altered plasma levels of soluble markers that potentially could reflect changes in amyloid processing was found in patients with persistent headache after SARS-CoV-2 vaccine and particular in those with persistent headache after COVID-19 with association to cognitive symptoms.

**How this study might affect research, practice or policy:** Our data underscore the need for more long-time follow-up of patients with new onset headache following COVID-19 or SARS-CoV-2 vaccination and this follow-up might also include blood tests for amyloid processing and neuroinflammation.

## Introduction

During the coronavirus disease (COVID)-19 pandemic, headache emerged as an important health issue, increasing the burden of headache disorders.^1, 2^ Both severe acute respiratory syndrome coronavirus 2 (SARS-CoV-2)) infection and, although to a lesser degree, SARS-CoV-2 vaccination, induced headache as a secondary headache disorder as well as negatively impact already existing primary headache disorder.^1, 3^ Those with primary headache disorder experience headache in the acute phase of COVID-19 and after vaccination more often than other SARS-CoV-2-infected or vaccinated individuals.^1^ Approximately one in five patients who present headache during the acute phase of COVID-19 get persistent headache, often with accompanying symptoms including cognitive symptoms and fatigue.^1^ Also, a subset of individuals with headache attributed to SARS-CoV-2 vaccinations develop long lasting headaches.^1, 3^ Furthermore, in patients with primary headache disorders, SARS-CoV-2 vaccination and in particular an acute SARS-CoV-2 infection worsened their chronic headache burden.^3^ The mechanisms for persistent headaches after SARS-CoV-2 infection or vaccines are, however, unknown. One proposed mechanism is persistent immune activation and inflammation triggered by SARS-CoV-2 or vaccination^4, 5^ with the trigeminovascular system as a potential target.^1^ Additionally, the inflammatory responses during acute COVID-19, involving the release of prostaglandins, interleukin (IL)-6 and other inflammatory cytokines could increase blood-brain barrier permeability leading to neuroinflammation with accompanying headache.^6, 7^

Serum Amyloid A (SAA1), an acute phase protein and systemic marker of inflammation, was early reported as a biomarker of poor prognosis in COVID-19.^8^ Recently, amyloid precursor protein (APP), a membrane protein considered to play a main role in Alzheimer disease pathology,^9^ was described as a new receptor for SARS-CoV-2 infection and revealed a previously unknown mechanistic insight into COVID-19 related neuropathological sequelae.^10^ Moreover, Pregnancy Zone Protein (PZP), originally known as a broad-spectrum immune-suppressive protein that suppresses T-cell function during pregnancy to prevent foetal rejection, has also been proposed as a biomarker for various inflammatory disorders,^11, 12^ including the prognosis of COVID-19.^13^ With relevance to CNS involvement, serum levels of PZP were higher in women with pre-symptomatic Alzheimer’s disease.^14^ Furthermore, Cathepsin L1 (CTSL), a lysosomal cysteine protease that plays a major role in intracellular protein catabolism, has been demonstrated to enhance SARS-CoV-2 virus entry, at least in pre-clinical models,^15^ and interestingly, CTSL has also been suggested to be involved in the degradation of certain amyloid fibrils.^16^ To this end, however, circulating levels of these biomarkers (i.e., SAA1, APP, PZP, and CTSL) in patients with persistent headaches after SARS-CoV-2 infection or vaccination, with or without headache during the acute phase of COVID-19 or immediately after vaccination, have not been reported.

In the present study, with the aim to give novel insight into the pathological mechanisms leading to persistent headache following COVID-19 and SARS-CoV-2 vaccination, we hypothesize that plasma levels of soluble markers that potentially could reflect changes in amyloid processing are related to persistent headache after SARS-CoV-2 vaccine and COVID-19 and particular in those with accompanying cognitive symptoms. This hypothesis was tested by measurement of relevant markers in plasma in patients with new onset persistent headache based on clinical assessments and validated questionnaires. The levels were compared in healthy controls that had undergone COVID-19 or SARS-CoV-2 vaccination but without headaches. The levels of these molecules were also assessed in relation to accompanying fatigue or cognitive symptoms, and in the COVID-19 group, the need for hospitalization during the acute infection.

## Methods

In this prospective observational cohort study performed at Norwegian neurological departments and led by Oslo University Hospital, Oslo, Norway, clinical characteristics and blood biomarker profiles were assessed in patients with severe headaches for a minimum of six months after COVID-19 or SARS-CoV-2 vaccination and compared to healthy controls. The inclusion period lasted from September 2020 to June 2023. We hypothesized that plasma levels of soluble markers that potentially could reflect changes in amyloid processing such as APP, PZP, CTSL, and SAA are linked to persistent headaches after SARS-CoV-2 vaccination and COVID-19 in those with accompanying cognitive symptoms. This hypothesis was tested by relevant plasma markers longitudinally patients with newly onset persistent headaches using clinical assessments and validated questionnaires. Differences in plasma levels were compared with healthy controls, and their relationship to cognitive symptoms was a major outcome.

### Study population and clinical assessments of the participants

The subgroup with **persistent headache after COVID-19 (n=29)** was recruited from the cohort of 149 individuals aged ≥ 18 years with persistent neurological symptoms 6 months after COVID-19 infection in the Norwegian observational multicentre study of nervous system manifestations and sequelae after COVID infection (NNC), a sub study of the ENERGY study led by European Academy of Neurology.^17^ The subgroup with **persistent headache after SARS-CoV-2 vaccine** (n=31) were participants in the Norwegian observational multi-centre study CovaxHEAD: New-onset severe headache after COVID-19 vaccine (NCT05235776) - a parallel study led by the same team as the NNC study collecting comparable variables.

Inclusion criteria for both subgroups were age ≥ 18 years and new-onset severe headache, or severe worsening of pre-existing headache, within one week after SARS-CoV-2 infection or vaccination. The degree of headache severity had to be so pronounced that the headaches were daily or almost daily and had resulted in significant disability or sick leave and still were the dominant symptom at 6-month follow-up. Exclusion criteria were short-lasting headaches, no follow-up assessments, or where other diagnoses were considered a better explanation for their symptoms (e.g. infections including COVID-19, head trauma, or somatoform disorders). Participants with COVID-19 who reported their headaches starting after SARS-CoV-2 vaccine were not included. None in this COVID-19 subgroup had SARS-CoV-2 vaccine induced headache although 24 participants (82.8%) had received SARS-CoV-2 vaccine. Types of vaccines were registered (BNT162b2 from BioNTech/Pfizer, mRNA-1273 from Moderna and ChAdOx1 nCoV-19 from AstraZeneca).

For both headache groups, an inclusion criterion was available plasma samples that could be used in biomarker analyses. CovaxHEAD included a total of 62 participants from 7 Norwegian centres. However, as blood samples for amyloid profiling were available only at Oslo University Hospital, only the participants included from Oslo University Hospital were included. In the NeuroCOVID, a total of 149 patients were included, with 46 (30%) categorised as having headache as the dominant symptom. However, in the present study, only participants with available samples for biomarker analyses were included, i.e., only patients from Oslo University Hospital were included in this biomarker substudy.

The cognitive symptoms were evaluated using questionnaires from the ENERGY study, led from the European Academy of Neurology.^17^ Additionally, all participants in the group with persistent headache after COVID-19 (n=29) were assessed with the Montreal Cognitive Assessment (MoCA), administered by a trained neurologist and defined as normal when MoCA score was above 26. The same questions were applied to the patients with chronic headaches following SARS-CoV-psychologist, neuropsychologist or psychiatrist as part of the multidisciplinary headache treatment.

The clinical data, including the headaches, were reviewed by a senior neurologist with headache expertise (AHA). The headaches were classified according to their phenotype. The clinical visits and first set of blood samples were collected at inclusion, mean 181.0 (SD 145.8) days after COVID-19 in the group with headache after COVID-19 and mean 290.3 (SD 145.8) days after SARS-CoV-2 vaccine in the group with headache after SARS-CoV-2 vaccine. Both groups had 6-month follow-up with clinical visits and blood samples thereafter.

Controls: individuals above 18 years of age, without any history migraine or other disabling headache disorders and who had never experienced neurological symptoms after SARS-CoV-2 vaccine or COVID-19. Furthermore, other exclusion criteria were anyimmunological disorders or use of immunosuppressive drugs. A total of 16 plasma samples were collected in the period April to July 2023 from 8 men and 8 women, mean age 40.6 years (SD 11.2). The healthy controls did not undergo detailed questionnaire or neurologic, neuropsychiatric or neuropsychological examination. Serum antibody assessments of the control group showed high levels in all participants (median 15831 BAU/ml, minimum 4309, maximum 64 676 BAU/ml) indicating strong or very strong immune response indicating that all had undergone COVID-19 or received SARS-CoV-2 vaccine.

### Blood sampling protocol

Peripheral venous blood was drawn into pyrogen-free blood collection tubes with ethylenediamine tetra acetic acid (EDTA) as an anticoagulant, immediately immersed in melting ice and centrifuged at 4°C 2500 × *g* for 20 minutes within 60 minutes to obtain platelet-poor plasma. The plasma samples were stored in multiple aliquots and thawed only once.

### Biomarker assessments

Plasma markers were assessed in all three groups – patients with persistent headache after COVID-19 infection (n=29), patients with persistent headache after the COVID-19 vaccine (n=31), and healthy controls (n=16). The participants had blood samples drawn when they were included in the study. Additionally, 6-month follow-up blood sampling was done in the headache groups. Plasma levels of APP (Cat# DY850), PZP (Cat# DY8280-05), CTSL (Cat# DY952), and SAA1 (Cat# DY3019-05) were measured by enzyme-linked immunosorbent assay (ELISA) using commercially available antibodies (R&D Systems, Minneapolis, MN) in a 384-format using a combination of a SELMA pipetting robot (Analytik Jena AG, Jena, Germany) and a BioTek dispenser/washer (BioTek Instruments, Winooski, VT). Absorption was read at 450 nm by using an EIA plate reader (BioTek Instruments) with wavelength correction set to 540 nm. Intra- and inter-assay coefficients of variation for these analyses were all <10%.

### Ethics approval

This study was conducted in accordance with the Helsinki Declaration as revised in 2013. Ethical approval for this study was obtained from The Norwegian South-Eastern Ethical Committee (152727 NNC, 351097 CovaxHEAD). Written informed consent was obtained from all subjects before inclusion into the study.

### Statistics

Statistical Package for Social Science (IBM SPSS Inc., version 29 for Windows) was used for statistical analysis. Differences in patient characteristics were tested using the Chi-square test or Fisher’s exact test for categorical variables, and the Mann-Whitney U test for continuous variables. Prevalence of characteristics (frequency and percentages) were calculated for each group. Plasma marker levels were compared between groups using a multivariate general linear model using markers as dependent, group as fixed, and age as covariate. A two-sided p-value of <0.05 was deemed statistically significant.

Patient and public involvement.

the user panel at the Department of Neurology, Oslo University Hospital and the user organization “Hodepine Norge” has contributed to the project. Hodepine Norge has been contributing to public meetings about the project.

## Results

### Characteristics and headache phenotypes in the study group

Demographics such as age, gender and co-morbidities are shown in Table 1. There was equal age and sex distribution in both headache groups (i.e., persistent headache following COVID-19 or SARS-CoV-2 vaccination, respectively) with 3 out of 4 participants being female. Comorbidity of frequent diseases, such as pre-existing migraine, allergies, anxiety, and depression were common in both groups. Few had cardiovascular or metabolic disorders. On third of the patients with persistent headache after COVID-19 infection was hospitalised in the acute phase of the infection. In the group with a persistent headache after SARS-CoV-2 vaccine 17 (54.8%) reported previous COVID-19 at baseline, but importantly, without any persistent headache after the infection (Table 1).

**Table 1.** Demographic and clinical characteristics at inclusion.

| Characteristics | Headaches<br>after<br>COVID-19<br>n = 29 | Headaches<br>SARS-CoV-2<br>vaccine<br>n = 31 | Control<br>group n=16 | P<br>value* |
| --- | --- | --- | --- | --- |
| Mean age at inclusion, (SD) | 43.5 (11.2) | 43.8 (12.4) | 40.6 (11.2) | 0.836 |
| Female, n (%) | 22 (75.9) | 24 (77.4) | 8 (50.0) | 0.862 |
| Education |  |  |  |  |
| Secondary school n (%) | 2 (6.8) | 7 (22.6) |  | 0.193 |
| College/university ≤ 4 years, n (%) | 14 (48.3) | 12 (38.7) |  |  |
| College/university > 4 years, n (%) | 13 (44.8) | 12 (38.7) |  |  |
| Hospitalisation during COVID-19, n (%) | 9 (31.0) | 3 (9.7) | 0 | 0.357 |
| Smoking, n (%) | 1 (4.4) | 2 (6.5) |  | 0.610 |
| Pre-existing migraine, n (%) | 11 (31.0) | 12 (38.7) | 0 | 0.871 |
| Comorbidities |  |  |  |  |
| Allergy, n (%) | 16 (55.1) | 13 (41.9) |  | 0.240 |
| Asthma, n (%) | 2 (6.9) | 6 (19.4) |  | 0.150 |
| Depression, n (%) | 8 (27.6) | 10 (32.3) |  | 0.759 |
| Anxiety, n (%) | 7 (24.1) | 13 (41.9) |  | 0.170 |
| Diabetes, n (%) | 0 | 1 (3.3) | 0 | 0.330 |
| Cardiovascular disease, n (%) | 0 | 1 (3.3) | 0 | 0.330 |
| Stroke, n (%) | 0 | 0 | 0 |  |
| Associated symptoms |  |  |  |  |
| Fatigue, n (%) | 22 (75.9) | 20 (64.5) |  | 0.204 |
| Subjective cognitive symptoms, n (%) | 22 (75.9) | 17 (54.8) |  | 0.119 |
| Sick leave at first visit, n (%) | 21 (72.4) | 18 (58.1) | . | 0.244 |
| Sick leave at 6-month follow-up, n (%) | 19 (65.5) | 14 (45.2) |  | 0.207 |

\*Comparison of the two headache groups.

Chronic migrainous headache was the dominant headache phenotype in both headache groups while tension-type headache was the second most common phenotype in both headache groups followed by episodic migrainous headache (Figure 1). Cranial neuralgia often accompanied by painful paraesthesia in the extremities was the dominant phenotype in 5 (17%) of the participants in the group with headaches after COVID-19.

**Figure 1.**
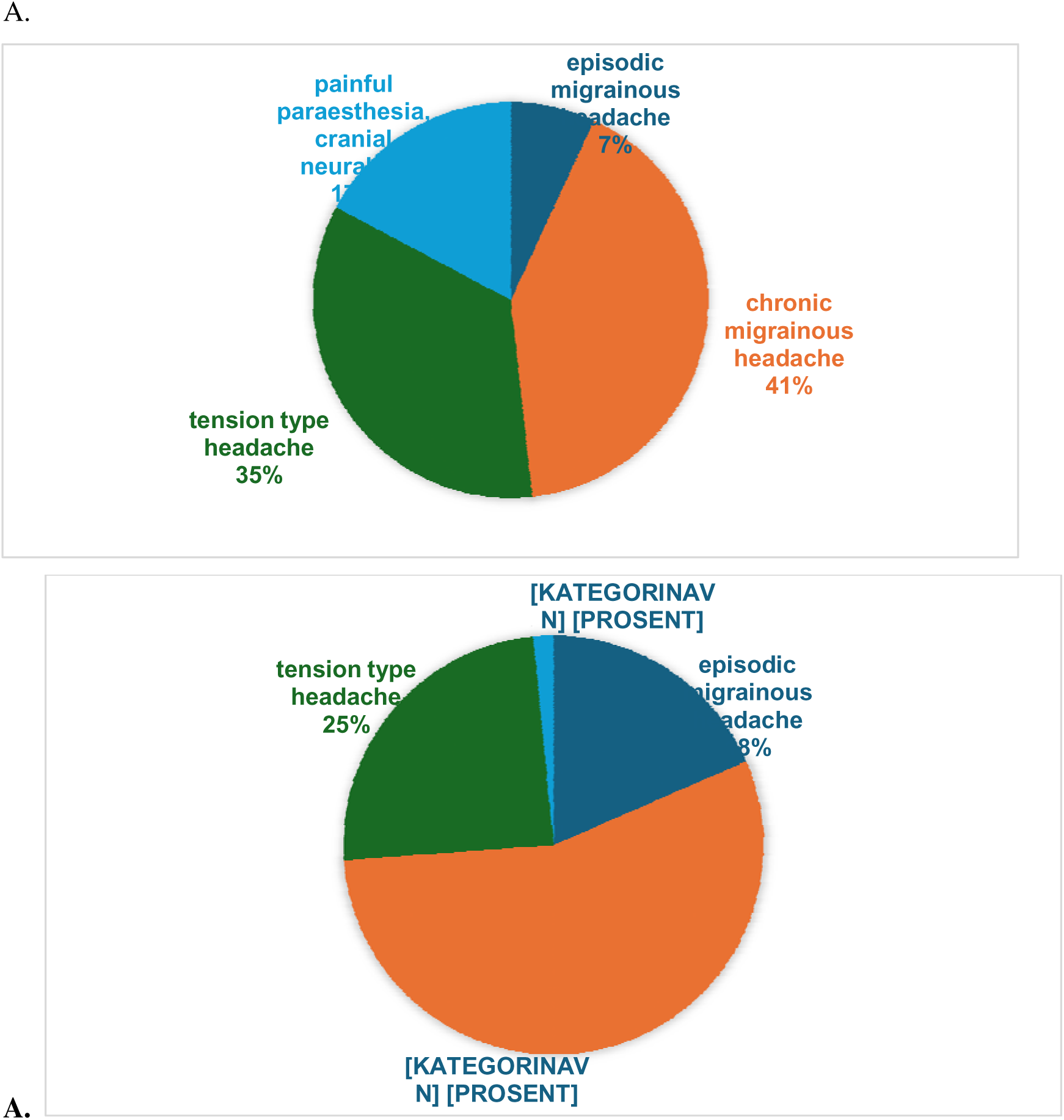
Phenotype of persistent headache reported after COVID-19 (A) and SARS-CoV-2 vaccine (B).

More than two thirds of the participants with headache post COVID-19 and more than half the participants with headache post SARS-CoV-2 vaccination were on sick-leave due to the current symptoms at inclusion with only a modest reduction in the percentages on sick-leave at 6-month follow-up (72.4 % to 65.5 % and 58.1 % to 45.2 %, post COVID-19 and SARS-CoV-2 vaccination, respectively; Table 1). Subjective cognitive symptoms and fatigue were common, but not dominating (see Methods), in the headache groups (Table 1). All the patients with COVID-19 had normal MoCA score except for one patient with critical illness myopathy in the acute phase of COVID-19 with a score of 24 at follow-up. The patients with chronic headaches after COVID-19 vaccine were assessed by a psychologist, neuropsychologist, or psychiatrist as part of the multidisciplinary headache treatment whereof three were diagnosed with depression, one with functional neurological disorder accompanying deterioration of chronic migraine, and two were not found to have any psychiatric disorders.

In the group with new onset headache after SARS-CoV-2 vaccine 34.5% reported the headache starting after their first vaccine, 41.4% after the second vaccine and 24.1% reported the headache to start after their third vaccine. Although the BNT162b2 vaccine was the most used vaccine, 89.7% reported their headache starting after RNA-1273 vaccine whereas 10.3% after BNT162b2 and none after ChAdOx1 nCoV-19.

### Biomarkers in relation to post-COVID-19 headache and headache following SARS-CoV-2 vaccination

At both inclusion and after 6 months, we found a strong and persistent upregulation of APP in patients with headache after COVID-19 as compared to individuals with vaccine induced headache and healthy controls (Figure 2A). In contrast, there was upregulation of CTSL in patients with headaches after vaccine compared to patients with headaches after COVID-19, but these differences were not seen at 6 months follow-up (Figure 2B). Furthermore, plasma levels of PZP were elevated in patients with headache after SARS-CoV-2 vaccination at both inclusion and after 6 months compared to controls, whereas a more modest increase was seen in those with COVID-19 associated headache only at inclusion (Figure 2C). As for SAA1, there were no differences between the two headache groups and healthy controls either at baseline or during follow-up (Figure 2D). Finally, whereas post-COVID-19 symptoms have been related to disease severity during the acute phase,^18^ we found no differences in the actual biomarkers between those who were hospitalized and those who were not hospitalized during the acute phase of COVID-19 (Supplemental Table 1).

**Figure 2.**
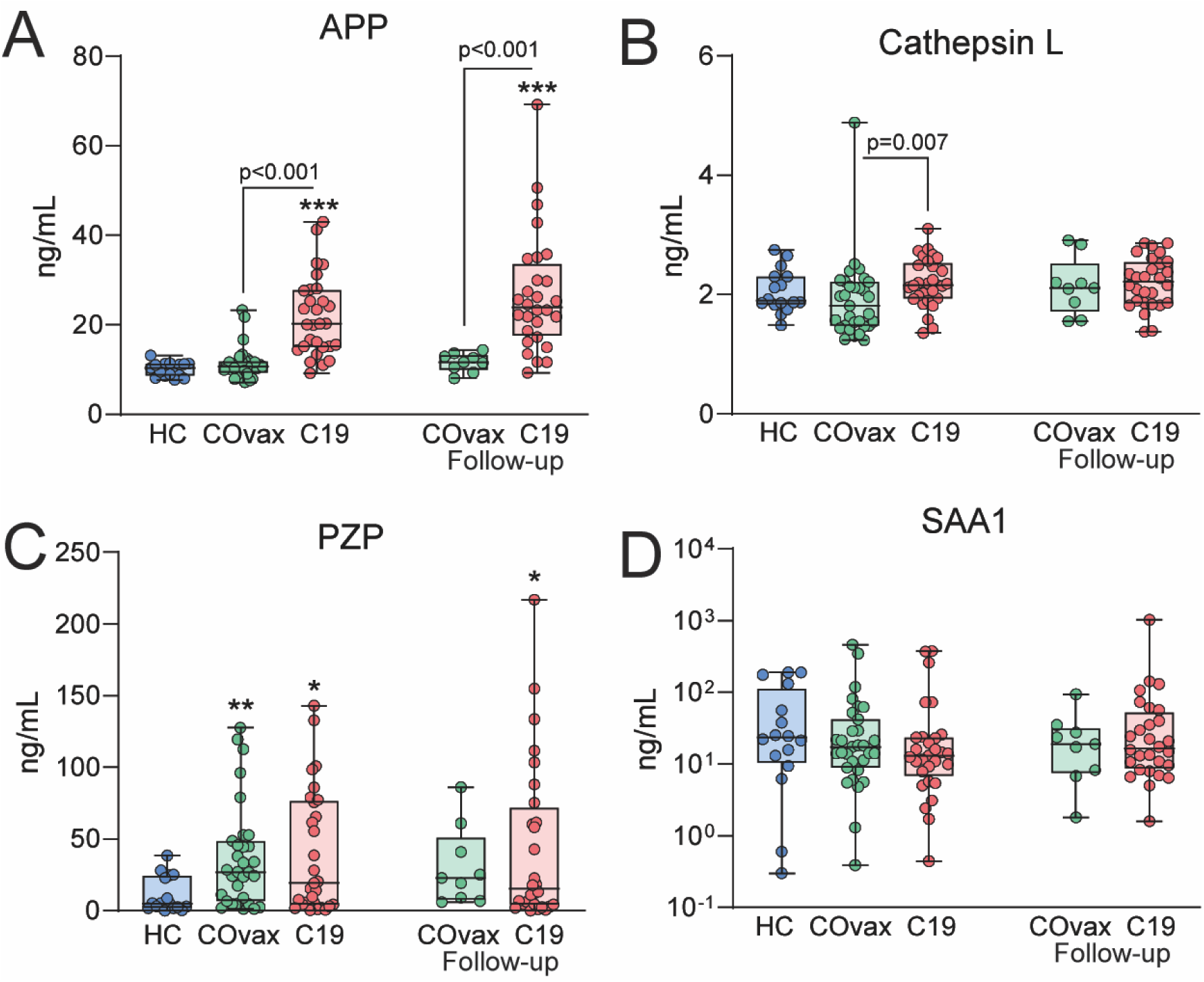
Plasma levels of **A.** Amyloid Precursor Protein (APP), **B**. Cathepsin L., **C.** Pregnancy Zone Protein (PZP) and **D**. Serum Amyloid A (SAA1) in healthy controls (HC) (n=16), participants with persistent headache after SarsCoV-2 vaccine (COvax) (n=31) and participants with persistent headache after COVID-19 (C19) (n=29).

### Biomarker levels in relation to associated fatigue and cognitive symptoms

There was a significant upregulation of APP among participants with accompanying cognitive symptoms in the group with persistent headache after COVID-19, but this pattern was not seen in those with headache after SARS-CoV-2 vaccination (Figure 3). Although those with fatigue also had higher levels of APP compared with those without, the difference did not reach statistical significance (Figure 3). As for PZP and CTSL, there were no differences between those with or without associated cognitive symptoms or fatigue, neither in the COVID-19 nor the SARS-CoV-2 group (Table 2).

**Figure 3.**
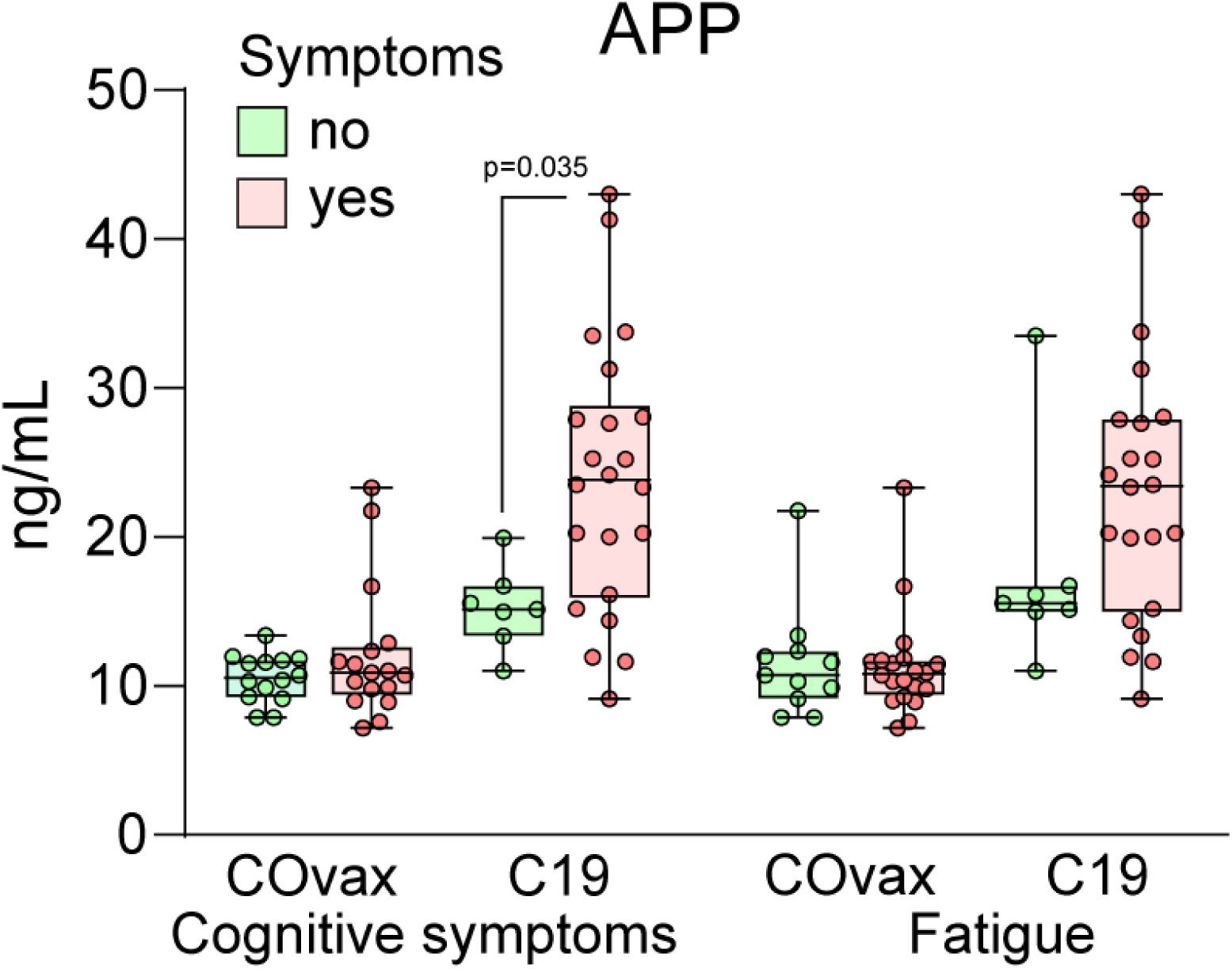
Plasma levels of Amyloid Precursor Protein (AAP) in relation to cognitive symptoms and fatigue in those with persistent headache after SARS-CoV-2 vaccine (COvax) or COVID-19 (C19).

**Table 2.**
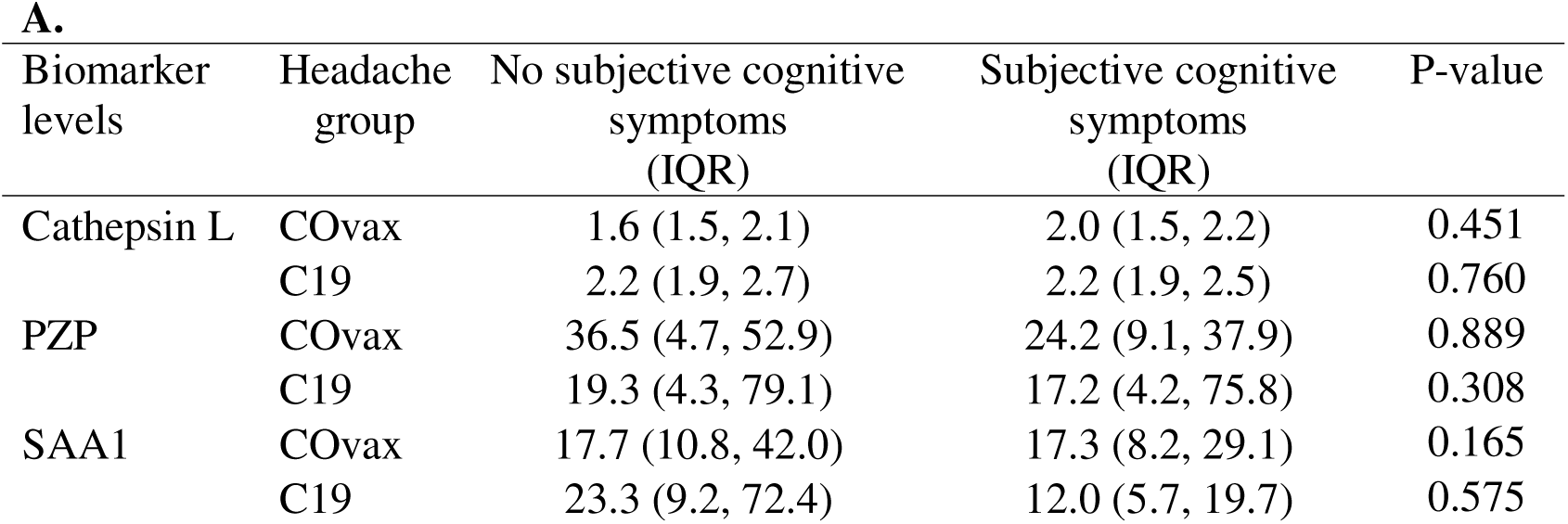
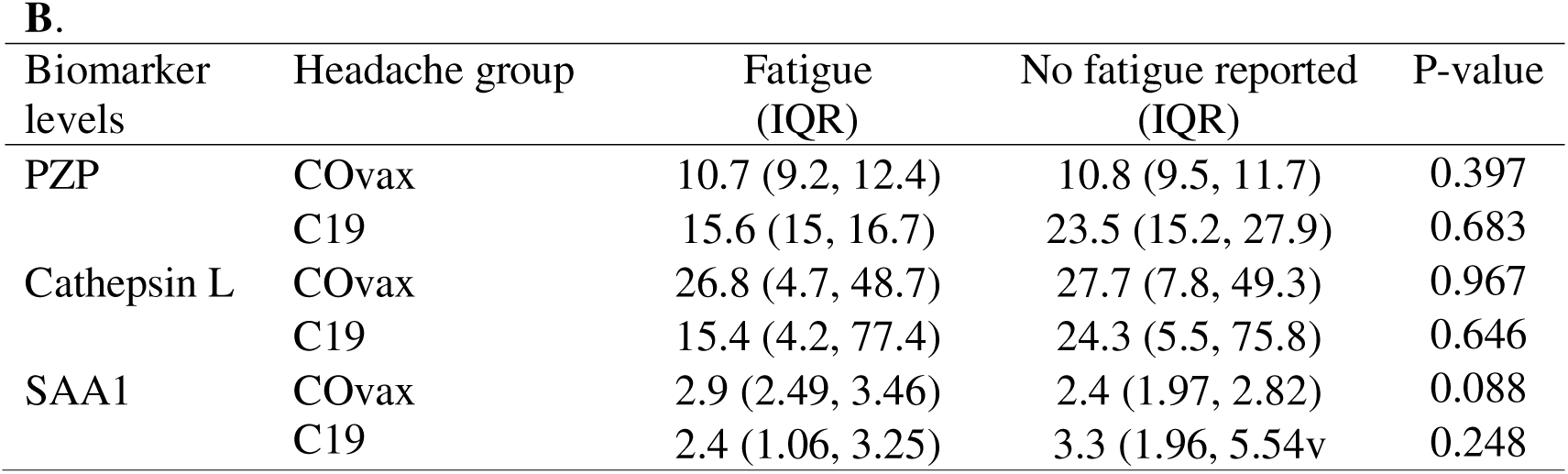
Median biomarker levels (ng/mL) with interquartile range (QR) of Cathepsin L, Pregnancy Zone Protein (PZP) and Serum Amyloid A (SAA1) in relation to **A.** subjective cognitive symptoms and **B.** fatigue in participants with persistent headache after SarsCoV-2 vaccine (COvax) (n=31) and participants with persistent headache after COVID-19 (C19) (n=29).

## Discussion

Persistent headache has been reported following acute COVID-19 and to some extent also after SARS-CoV-2 vaccination.^1, 3^ In the present study, we observed higher plasma levels of APP in patients with persistent headache after COVID-19, but not in patients with persistent headache following SARS-CoV-2 vaccination, compared to healthy controls. Remarkably, elevated APP levels persisted at 6-months follow-up and was associated with accompanying cognitive symptoms. Notably, plasma levels of CTSL were higher in patients with headaches after SARS-CoV-2 vaccination compared to those with headaches after COVID-19.

Furthermore, PZP levels were persistently elevated in patients with headache after SARS-CoV-2 vaccination, while a more modest increase was observed in those with COVID-19 associated headache as compared to healthy controls. To the best of our knowledge, this is the first report linking proteins in amyloid formation to persistent headache following COVID-19, and CTSL and PZP to persistent headache after SARS-CoV-2 vaccination.

As depicted in Figure 4, our data point to plausible mechanism of serum APP and PZP proteins in relation to COVID-19 and SARS-CoV-2 vaccine induced headache. In brief, APP is produced by neurons, and proteolysis by various secretases can generate a soluble (s) fragment, sAPP, as well as release the neurotoxic Aβ peptide. SARS-CoV-2 infection may increase APP expression via the ACE2 receptor^19, 20^ and APP promotes cellular entry of the virus enhancing Aβ-associated pathology.^10^ The Aβ peptide may accumulate to form amyloid plaques^21^, leading to activation of astrocytes and microglia, and promote neuroinflammation by different mechanisms^22^ with potentially affecting headache symptoms.^23^

**Figure 4.**
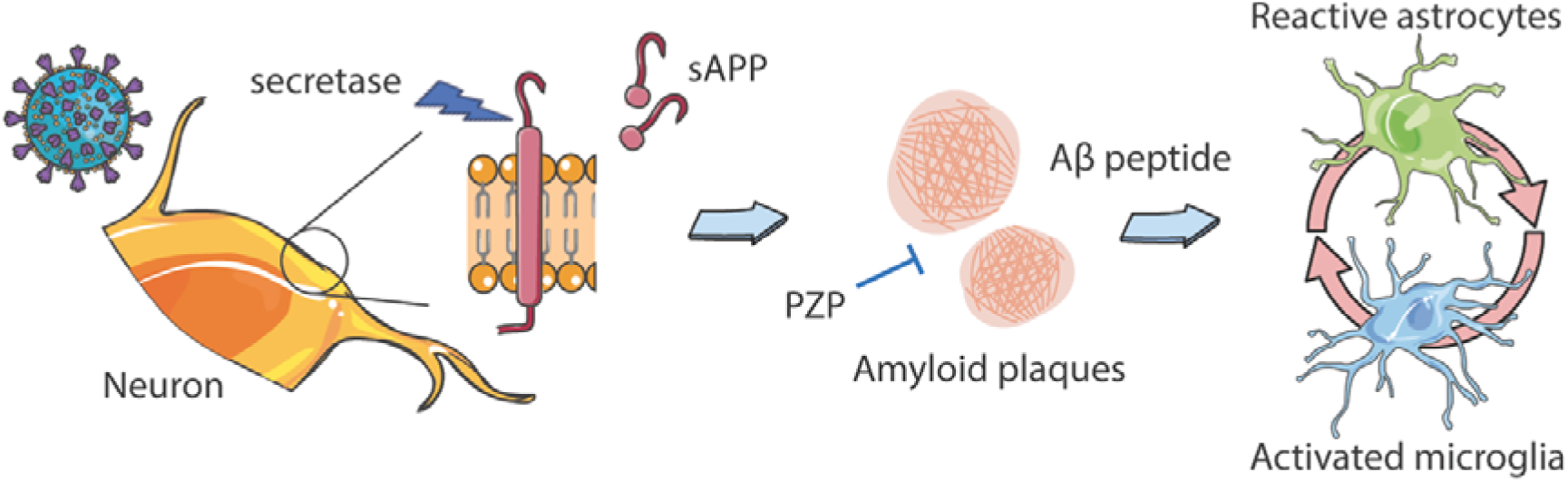
Possible mechanisms of soluble Amyloid Precursor Protein (sAPP) and Pregnancy Zone Protein (PZP) in relation to COVID-19 and SARS-CoV-2 vaccination associated headache. The authors wish to acknowledge SERVIER Medical Art (www.servier.fr) for use of their medical art kits when making the illustration in the article.

Increased levels of APP are reported as marker for preclinical Alzheimer disease; however, the biological function of this protein extends beyond amyloid formation^24^ and elevated levels have been found in various metabolic and cardiovascular disorders.^25^ Like in some other viruses, amyloids are identified in the SARS-CoV-2 proteins, i.e., the structural spike, the nucleocapsid, and the accessory ORF6/ORF10 proteins and this may also be of clinical relevance.^26^ There are several reports on the worsening of preexisting dementia including Alzheimer’s Disease and the development of cognitive impairment following SARS-CoV-2 infection, but the pathogenesis of these changes is still incompletely understood.^26-28^ However, in a recent study on the relationship between COVID-19 and Alzheimer’s disease the changes in blood biomarkers were linked to brain structural imaging patterns associated with Alzheimer’s disease, lower cognitive test scores, and poorer overall health evaluations.^26-29^

In the present study few or none had such clinically verified co-morbidities. Still, we found significant upregulation of APP among participants with accompanying cognitive symptoms in the group with persistent headache after COVID-19 compared to those without cognitive symptoms. Moreover, Camacho et al. reporting network analyses of SARS-CoV-2 infection in brain microvascular endothelial cells suggested increased expression of APP.^19^ In a previous study of patients with COVID-19 associated neurological syndromes, signs of impaired amyloid processing were found with significantly reduced levels of soluble amyloid precursor protein in CSF from patients compared to controls.^30^ However, the study primarily included patients with severe neurological diseases such as Guillain Barre syndrome and encephalopathy and only one patient was described with central pain syndrome.^30^ Furthermore, cognitive symptoms were not reported. At present, however, the reason for as well as the consequence of the raised APP in patients with persistent headache following COVID-19 is elusive and follow-up studies are clearly needed.

Recent studies suggest that serum markers, known to be associated with cognitive impairment when measured in CSF, are also linked to cognitive impairment.^31^ Thus, Urbano T et al. found a positive correlation between neurofilament light chain protein levels in serum and CSF in patients with neurocognitive disorders.^31^ Moreover, a recent study revealed that the changes in blood biomarkers were linked to brain structural imaging patterns associated with Alzheimer’s disease, as well as and lower cognitive test scores. Several of these proteins such as Tau, are known to be associated with Alzheimer’s disease when detected in CSF.^29^ To the best of our knowledge, whether a similar pattern will be seen for APP and PZP has not been studied.

Although a modest increase in the COVID-19 related headache group at inclusion, plasma levels of PZP were persistently elevated only those with prolonged headache following SARS-CoV-2 vaccination. The reason for this pattern remains unclear. PZP is known to promote a beneficial immunosuppression during pregnancy to prevent rejection of the foetus. More recently, increased PZP levels have been observed during airway infections,^12^ and have been suggested as a marker of certain cancers.^32^ Moreover, previous studies with mouse models have indicated that APP may increase the entry of the SARS-Cov-2 virus into cells, while PZP has been described to inhibiting amyloid aggregation.^33^ Since APP was lower in the vaccine group than the COVID-19 group, one intriguing hypothesis is that PZP may have inhibited amyloid aggregation to a higher extent in the vaccine group than in the COVID-19 group. However, PZP has also been reported to stabilize misfolded proteins like amyloid β protein, and its role in amyloid formation remains unclear.^33^ Based on this relatively small study, which is at least hypothesis generating, it is hard to draw any firm conclusion regarding primary and secondary effects. In our opinion data in the literature regarding APP are more convincing in relation to its role in cognitive impairment compared to PZP. Therefore, we currently consider elevated APP as the most important finding, with potential some “additive” effects from PZP.

Since one of the three vaccines was disproportionately associated with headache, it warrants a discussion of potential mechanisms. While the ChAdOx1 nCoV-19 vaccine has been associated with severe cerebral events,^34^ none of the patients in the present study experienced new-onset persistent headaches after ChAdOx1 nCoV-19 vaccines. Moreover, although the BNT162b2 vaccine was the most commonly vaccine, 89.7% of the participants reported that their headache started after RNA-1273 vaccine. The reason for this pattern is not clear, but interestingly, an online survey on self-reported adverse reactions in individuals who had received two doses of either the BNT162b2 or mRNA-1273 vaccine reported more side effects among those receiving the mRNA-1273 vaccine.^35^ One could speculate that it was related to the higher mRNA content in mRNA-1273 as compared with the BNT162b2 vaccine.^36^ However, based on the low number of patients in the present study, the results should be interpreted with caution.

The strengths of this study were the comprehensive examinations of the participants done by headache specialists with precise phenotyping of the headache syndromes and the strong multidisciplinary collaboration. The major limitations were the lack of CSF samples in both headache groups and small sample size. Moreover, we lack parallel samples from CSF and measurements of the actual protein in individuals that have undergone COVID-19 and SARS-CoV-2 vaccination without the development of persistent headache. In particular, the control group was small and could have been matched better in relation to age and undergone longitudinal testing and been interviewed with more clinical information as in the patient groups.

In summary, altered plasma levels of soluble markers that potentially could reflect changes in amyloid processing was found in patients with persistent headache after SARS-CoV-2 vaccine and particular in those with persistent headache after COVID-19 with association to cognitive symptoms. Future and larger studies that also include CSF samples are needed to further elaborate the clinical consequences of these findings.

## Data Availability

All data produced in the present study are available upon reasonable request to the authors

## Acknowledgement

We thank Marcus Boateng for registering data and Katrine Persgård Lund and Azita Rashidi with the sampling of blood biomarkers.

## Statistical Analysis

Thor Ueland and Anne Hege Aamodt

---

**Supplemental Table 1.** Plasma levels (ng/mL) of Amyloid Precursor Protein (APP), Cathepsin L, Pregnancy Zone Protein (PZP) and Serum Amyloid A (SAA1) in participants with persistent headache after COVID-19 (C19) in relation to severity of the acute infection (n=29).

| **Test statistics** | **Biomarkers** |  |  |  |
| --- | --- | --- | --- | --- |
|  | **APP** | **Cathepsin L** | **PZP** | **SAA1** |
| Mann-Whitney U | 61.00 | 49.00 | 60.00 | 69.00 |
| Wilcoxon W | 97.00 | 280.0 | 96.00 | 96.00 |
| Z | -1,12 | -1.71 | -1.17 | -0.73 |
| Asymptotic Sig. (2-tailed) | 0.26 | 0.09 | 0.24 | 0.46 |
| Exact Sig | 0.28 | 0.93 | 0.26 | 0.49 |

**Supplemental Table 2.**

| Co-investigators | Affiliations |
| --- | --- |
| Øyvind Torkildsen | Neuro-SysMed, Department of Neurology, Haukeland University Hospital, Bergen, Norway; Department of Clinical Medicine, University of Bergen, Bergen, Norway |
| Vojtech Novotny | Department of Neurology, Haukeland University Hospital, Bergen, Norway |
| Heidi Øyen Flemmen | Department of Neurology, Telemark Hospital Trust, Skien, Norway |
| Hilde Karen Ofte | Department of Neurology, Nordlandssykehuset, Bodø, Norway |
| Christian Samsonsen | Department of Neurology of Clinical Neurophysiology, St. Olav Hospital, Trondheim, Norway |
| Barbara Ratajczak-Tretel | Department of Neurology, Østfold Hospital Trust, Sarpsborg, Norway |
| Anette Huuse Farmen | Department of Neurology, Innlandet Hospital Trust, Lillehammer, Norway |
| Per Selnes | Department of Neurology, Akershus University Hospital, Nordbyhagen, Norway and Institute of Clinical Medicine, Faculty of Medicine, University of Oslo, Oslo, Norway |
| Stein Andersson | Department of Psychology, University of Oslo, Oslo, Norway |
| Erlend Bøen | Unit for Psychosomatics and C-L psychiatry for adults, Oslo University Hospital, Oslo, Norway |
| Siv Elin Pignatiello | Unit for Psychosomatics and C-L psychiatry for adults, Oslo University Hospital, Oslo, Norway and Institute of Clinical Medicine, Faculty of Medicine, University of Oslo, Oslo, Norway |

## References

1. Caronna E, van den Hoek TC, Bolay H, et al. Headache attributed to SARS-CoV-2 infection, vaccination and the impact on primary headache disorders of the COVID-19 pandemic: A comprehensive review. Cephalalgia 2023;43:3331024221131337.

2. Shen Q, Joyce EE, Ebrahimi OV, et al. COVID-19 illness severity and 2-year prevalence of physical symptoms: an observational study in Iceland, Sweden, Norway and Denmark. Lancet Reg Health Eur 2023;35:100756.

3. Atalar AC, Acarli ANO, Baykan B, et al. COVID-19 vaccination-related headache showed two different clusters in the long-term course: a prospective multicenter follow-up study (COVA-Head Study). J Headache Pain 2023;24:132.

4. Files JK, Sarkar S, Fram TR, et al. Duration of post-COVID-19 symptoms is associated with sustained SARS-CoV-2-specific immune responses. JCI Insight 2021;6.

5. Gaebler C, Wang Z, Lorenzi JCC, et al. Evolution of antibody immunity to SARS-CoV-2. Nature 2021;591:639–644.

6. Garces KN, Cocores AN, Goadsby PJ, Monteith TS. Headache After Vaccination: An Update on Recent Clinical Trials and Real-World Reporting. Curr Pain Headache Rep 2022;26:895–918.

7. Hervé CL, B.; Del Giudice, G.; Didierlaurent, AM.; Tavares Da Silva, F.. The how’s and what’s of vaccine reactogenicity. NPJ Vaccines 2019;4.

8. Li HX, X.; Ren, H.; Xu, L.; Zhao, L.; Chen, X.; Long, H.; Wang, Q.; Wua, Q. Serum Amyloid A is a biomarker of severe Coronavirus Disease and poor prognosis. J Infect 2020;80:646–655.

9. Orobets KS, Karamyshev AL. Amyloid Precursor Protein and Alzheimer’s Disease. Int J Mol Sci 2023;24.

10. Chen J, Chen J, Lei Z, et al. Amyloid precursor protein facilitates SARS-CoV-2 virus entry into cells and enhances amyloid-beta-associated pathology in APP/PS1 mouse model of Alzheimer’s disease. Transl Psychiatry 2023;13:396.

11. Shao J, Jin Y, Shao C, Fan H, Wang X, Yang G. Serum exosomal pregnancy zone protein as a promising biomarker in inflammatory bowel disease. Cell Mol Biol Lett 2021;26:36.

12. Finch S, Shoemark A, Dicker AJ, et al. Pregnancy Zone Protein Is Associated with Airway Infection, Neutrophil Extracellular Trap Formation, and Disease Severity in Bronchiectasis. Am J Respir Crit Care Med 2019;200:992–1001.

13. Villar M, Urra JM, Rodriguez-Del-Rio FJ, et al. Characterization by Quantitative Serum Proteomics of Immune-Related Prognostic Biomarkers for COVID-19 Symptomatology. Front Immunol 2021;12:730710.

14. Ijsselstijn L, Dekker LJ, Stingl C, et al. Serum levels of pregnancy zone protein are elevated in presymptomatic Alzheimer’s disease. J Proteome Res 2011;10:4902–4910.

15. Zhao MM, Yang WL, Yang FY, et al. Cathepsin L plays a key role in SARS-CoV-2 infection in humans and humanized mice and is a promising target for new drug development. Signal Transduct Target Ther 2021;6:134.

16. Wang H, Sang N, Zhang C, Raghupathi R, Tanzi RE, Saunders A. Cathepsin L Mediates the Degradation of Novel APP C-Terminal Fragments. Biochemistry 2015;54:2806–2816.

17. Beghi E, Moro E, Davidescu EI, et al. Comparative features and outcomes of major neurological complications of COVID-19. Eur J Neurol 2023;30:413–433.

18. Headache Classification Committee of the International Headache Society (IHS) The International Classification of Headache Disorders, 3rd edition. Cephalalgia 2018;38:1-211.

19. Camacho RC, Alabed S, Zhou H, Chang SL. Network Meta-analysis on the Changes of Amyloid Precursor Protein Expression Following SARS-CoV-2 Infection. J Neuroimmune Pharmacol 2021;16:756–769.

20. Caradonna A, Patel T, Toleska M, Alabed S, Chang SL. Meta-Analysis of APP Expression Modulated by SARS-CoV-2 Infection via the ACE2 Receptor. Int J Mol Sci 2022;23.

21. O’Brien RJ, Wong PC. Amyloid precursor protein processing and Alzheimer’s disease. Annu Rev Neurosci 2011;34:185–204.

22. Di Benedetto G, Burgaletto C, Bellanca CM, Munafo A, Bernardini R, Cantarella G. Role of Microglia and Astrocytes in Alzheimer’s Disease: From Neuroinflammation to Ca(2+) Homeostasis Dysregulation. Cells 2022;11.

23. Kursun O, Yemisci M, van den Maagdenberg A, Karatas H. Migraine and neuroinflammation: the inflammasome perspective. J Headache Pain 2021;22:55.

24. Zheng H, Koo EH. The amyloid precursor protein: beyond amyloid. Mol Neurodegener 2006;1:5.

25. Guo Y, Wang Q, Chen S, Xu C. Functions of amyloid precursor protein in metabolic diseases. Metabolism 2021;115:154454.

26. Navolokin N, Adushkina V, Zlatogorskaya D, et al. Promising Strategies to Reduce the SARS-CoV-2 Amyloid Deposition in the Brain and Prevent COVID-19-Exacerbated Dementia and Alzheimer’s Disease. Pharmaceuticals (Basel) 2024;17.

27. Danta CC. Calcium Channel Blockers: A Possible Potential Therapeutic Strategy for the Treatment of Alzheimer’s Dementia Patients with SARS-CoV-2 Infection. ACS Chem Neurosci 2020;11:2145–2148.

28. Zhou Y, Xu J, Hou Y, et al. Network medicine links SARS-CoV-2/COVID-19 infection to brain microvascular injury and neuroinflammation in dementia-like cognitive impairment. Alzheimers Res Ther 2021;13:110.

29. Duff EP, Zetterberg H, Heslegrave A, et al. Plasma proteomic evidence for increased beta-amyloid pathology after SARS-CoV-2 infection. Nat Med 2025;31:797–806.

30. Ziff OJ, Ashton NJ, Mehta PR, et al. Amyloid processing in COVID-19-associated neurological syndromes. J Neurochem 2022;161:146–157.

31. Urbano T, Maramotti R, Tondelli M, et al. Comparison of Serum and Cerebrospinal Fluid Neurofilament Light Chain Concentrations Measured by Ella and Lumipulse in Patients with Cognitive Impairment. Diagnostics (Basel) 2024;14.

32. Huang J, Xu Y, Chen Y, et al. Revisiting the role of pregnancy zone protein (PZP) as a cancer biomarker in the immunotherapy era. J Transl Med 2024;22:500.

33. Cater JH, Kumita JR, Zeineddine Abdallah R, et al. Human pregnancy zone protein stabilizes misfolded proteins including preeclampsia- and Alzheimer’s-associated amyloid beta peptide. Proc Natl Acad Sci U S A 2019;116:6101–6110.

34. Schultz NH, Sorvoll IH, Michelsen AE, et al. Thrombosis and Thrombocytopenia after ChAdOx1 nCoV-19 Vaccination. N Engl J Med 2021;384:2124–2130.

35. Kitagawa H, Kaiki Y, Sugiyama A, et al. Adverse reactions to the BNT162b2 and mRNA-1273 mRNA COVID-19 vaccines in Japan. J Infect Chemother 2022;28:576–581.

36. Dickerman BA, Gerlovin H, Madenci AL, et al. Comparative Effectiveness of BNT162b2 and mRNA-1273 Vaccines in U.S. Veterans. N Engl J Med 2022;386:105–115.

